# Within-cohort discrimination and cross-cohort transfer of EEG-based ADHD classifiers: An exploratory two-cohort evaluation

**DOI:** 10.64898/2026.10.01.26364474

**Authors:** Buchan Liang

## Abstract

We evaluated whether participant-separated within-cohort discrimination was retained when electroencephalography (EEG) classifiers for attention-deficit/hyperactivity disorder (ADHD) were applied to a different population and recording task. This exploratory reanalysis included 121 children performing a visual task and 79 adults recorded during eyes-open rest after basic channel screening. Both cohorts used Cz/F4, 128 Hz, and the first fifteen non-overlapping 2-second epochs per participant. Fixed logistic regression and random forest classifiers used spectral, amplitude-derived, and combined features. Internal evaluation used five repetitions of five-fold participant-grouped cross-validation; bidirectional transfer fitted transformations and classifiers only on the source cohort. Adult spectral balanced accuracy (BA) was 0.9519–0.9546, compared with 0.5504–0.5664 in children. Adult spectral logistic regression exceeded all 999 internal label permutations (Monte Carlo p=0.001; within-family Holm-adjusted p=0.002). Paired spectral-minus-amplitude conditional intervals excluded zero in adults but spanned zero in children. Cross-cohort spectral BA was 0.3584–0.4675, with area under the receiver operating characteristic curve (AUC) of 0.0309–0.3857 in the unchanged score direction. Increasing pediatric source-epoch budgets moved random-forest AUC toward 0.5, whereas logistic-regression AUC remained below 0.1. Strong adult internal discrimination thus coexisted with reverse cross-cohort ranking. These findings characterize the evaluated pipelines, not clinical diagnostic utility or a unique mechanism: age, task, and acquisition differences were not separated, target results had previously been inspected, and conditional intervals excluded model-refitting uncertainty.

## Introduction

EEG classification for ADHD requires distinguishing prediction in the development population from prediction in a different recording cohort. Participant-separated validation prevents an individual’s epochs from appearing in both training and testing, but the remaining participants can still share recruitment, task, and acquisition characteristics. Cross-cohort evaluation asks whether a source-fitted scoring rule remains useful outside that setting. Here, robustness refers specifically to this evaluated transfer behavior and its sensitivity to the source-recording budget, not to resistance to every form of noise or distribution shift.

Independent EEG–ADHD validation has a substantial history. Snyder et al. [1] evaluated an EEG-based approach in a blinded, multicenter clinical sample. Mueller et al. [2] developed an event-related-potential classifier in 75 adults with ADHD and 75 controls, then tested 17 independent ADHD participants. The reported 94% patient identification could not estimate external specificity because the additional sample contained no controls. Helgadóttir et al. [3] reported 76% accuracy in an independent cohort using coherence and age. These studies distinguish independent patient detection, two-class external discrimination, and internal cross-validation; their percentages do not represent interchangeable measures.

Validation design also affects reported performance. Brookshire et al. [4] demonstrated inflated EEG estimates when segments from the same participant appeared in both training and testing. Pulini et al. [5] examined sample size, circular analysis, and reported accuracy in ADHD neuroimaging. In an ADHD–EEG example, MRS-SHAP reported 98.40% segment-level accuracy, approximately 79.5% under grouped validation, and approximately 65.7% in a participant holdout [6]. High scores have also been reported under subject-separated designs, as in the ADHDeepNet preprint [7]. These studies motivate explicit reporting of validation units, but differences in metrics, montages, preprocessing, and models preclude a matched performance comparison with the present analysis.

Population and recording context introduce additional distinctions. Poil et al. [8] investigated age-dependent EEG differences; Kitsune et al. [9] showed that recording context within a session can change ADHD-related spectral contrasts. Dubreuil-Vall et al. [10] evaluated adult ADHD classification using task-related and resting inputs. These findings make age and task plausible contributors to cross-cohort differences without identifying the cause of any particular transfer result.

We therefore examined two public cohorts under one restricted processing specification: a pediatric visual-task cohort and an adult eyes-open resting-state cohort. The analysis addresses three questions: how well supplied diagnostic labels can be discriminated within each cohort; how spectral models compare with amplitude-derived models on identical participant splits; and how source-fitted models perform in the other cohort without adaptation. Bidirectional evaluation and a source-epoch budget series make the dependence on transfer direction and signal use explicit. Common channels, sampling rate, and duration reduce selected processing differences without equating the acquisition settings. The contribution is a traceable empirical evaluation, not a new classifier architecture or a matched replication of published high-accuracy models. S1 File provides the related-work comparison and separates source descriptions from unresolved methodological details.

## Materials and methods

### Study design and analysis record

This was an exploratory secondary analysis. Results from both cohorts had been inspected before the common-processing specification was fixed; source-only fitting therefore describes the evaluated runs, not a prospectively blinded external-validation design. The frozen configuration is `public_matched_processing_review_v3`. We report both classifiers and all three feature sets without a model-selection search within this matrix. Permutation analyses were restricted to the spectral models specified in the subsection “Uncertainty and permutation analyses”. Analyses of other datasets are outside the study scope.

### Cohorts and quality screening

The pediatric cohort (N) comprised 121 participants: 61 with ADHD labels and 60 controls. The supplied CSV mirror is associated with the Nasrabadi/IEEE DataPort resource, DOI 10.21227/rzfh-zn36, and contains 19 named channels sampled at 128 Hz during a visual task [11, 12]. The archived conversion check reconstructed 8,400 complete 2-second epochs from 2,166,383 CSV rows and matched the supplied epoch archive exactly. This checks the mirror-to-epoch conversion, not equivalence to the canonical IEEE files. The mirror’s upstream version and raw-file equivalence remain unresolved. Clinical labels were used as supplied. Individual ages, medication information, voltage units, and acquisition preprocessing were not independently verified in this reanalysis.

The adult cohort (M) used Mendeley Data version 1, DOI 10.17632/6k4g25fhzg.1 [13]. Its four MAT containers, organized by recorded sex and diagnosis, hold 80 records: 38 ADHD and 42 controls, with 56 male and 24 female records. Each MAT file contains only one diagnostic class, and diagnosis can therefore be recovered from file identity. This storage organization does not establish that the files were acquired or processed under different conditions. Any unmeasured acquisition or processing differences aligned with the diagnosis-defined file groups, if present, would remain unresolved by the available metadata. The primary analysis used only `cell1`, documented as 30 seconds of eyes-open rest recorded at Cz/F4 and 256 Hz. A record had to have shape 7680×2, finite samples, nonzero peak-to-peak range, and more than two distinct values in each required channel. One constant-channel record was excluded, leaving 79 participants: 37 ADHD and 42 controls, with 56 male and 23 female participants. These are basic signal-validity checks rather than comprehensive physiological artifact rejection.

Electrode identities were assigned from the task dictionary, not inferred from the number of array columns. Later adult task cells were checked for equality of array shape, data type, and decoded C-order sample bytes. They were not pooled into the primary analysis. The cohort characteristics, input hashes, task dictionary, and provenance limitations are documented in S2 File.

### Common signal processing

Cz and F4 formed the shared pair between the pediatric montage and the documented adult cell1 recording. Both cohorts were restricted to this pair in that order, with pediatric channel-name assertions. Adult records were resampled independently with `scipy.signal.resample_poly(up=1, down=2)` and divided into fifteen consecutive, non-overlapping 256-sample epochs. The pediatric analysis selected the first fifteen existing 2-second epochs per participant. Thus each person contributed 30 seconds in both source and target roles: 1,815 pediatric epochs and 1,185 adult epochs (Fig 1). These are repeated observations from 121 and 79 participants, respectively, not 3,000 independent participants. Epochs were the fitting observations; participants were the splitting, aggregation, evaluation, and resampling units.

**Fig 1.**
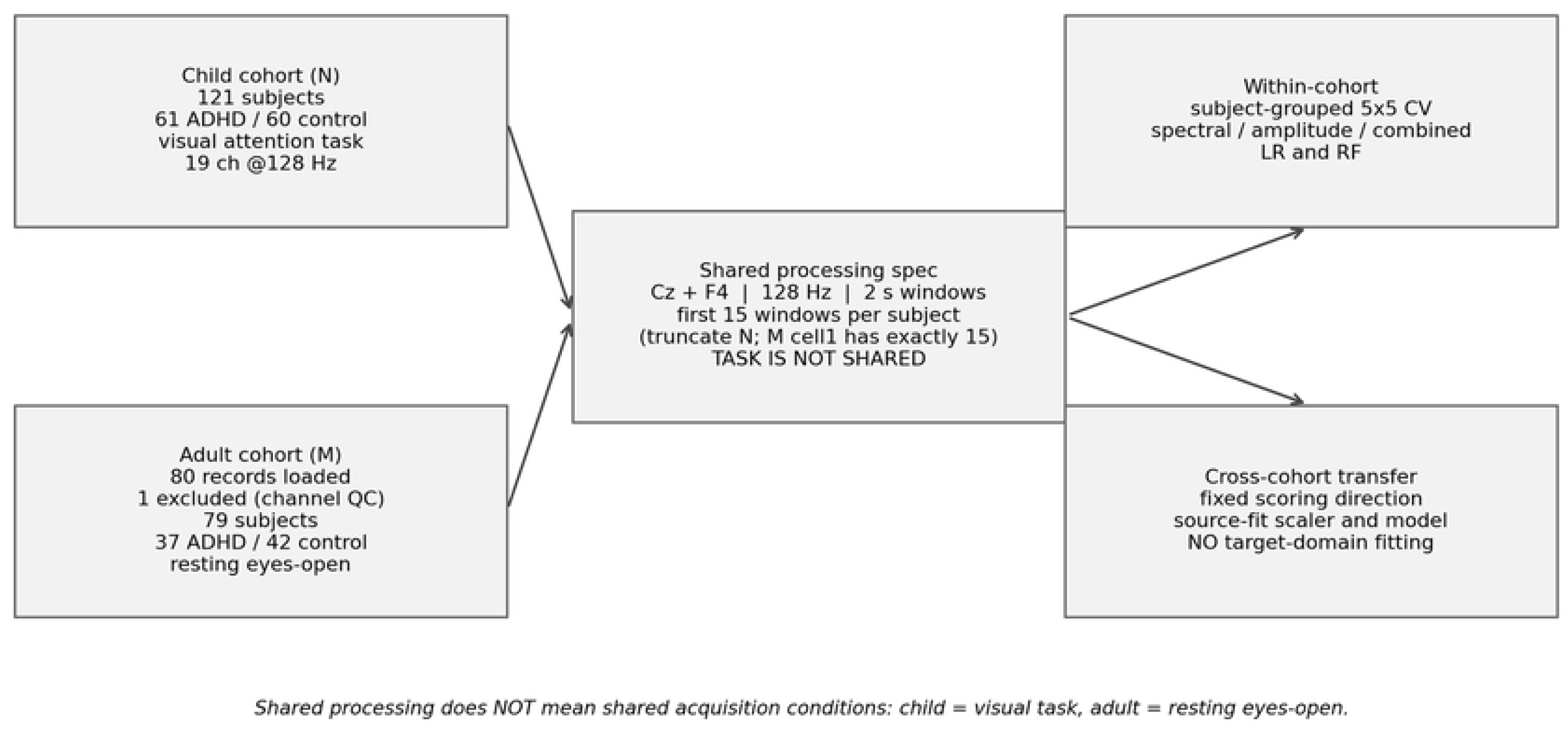
Data and evaluation design. The adult cell1 cohort retains 79 of 80 records after basic channel screening; all 121 pediatric participants meet the temporal-budget requirement. Both cohorts use Cz/F4, 128 Hz, and fifteen consecutive 2-second epochs per participant. Internal validation refits within training folds; transfer refits using the entire selected source cohort and evaluates the other cohort without target fitting. The pediatric visual task and adult eyes-open rest remain different acquisition contexts. AI-assisted drafting and integration of this workflow diagram used ChatGPT (OpenAI) and OpenAI Codex; exact historical model versions could not be recovered. The diagram content was checked against the frozen analysis configuration.

Adult resampling used float64 input, followed by float32 epoch storage. No additional independent-component analysis, notch filter, re-referencing, or dedicated band-pass filter was applied. The 1–45 Hz normalization range below is a feature definition, not a band-pass operation. Common channels, sampling rate, and temporal budgets standardize selected processing choices; they do not align the pediatric visual task with adult eyes-open rest or establish equivalent acquisition conditions.

### Feature sets and classifiers

The spectral set comprised 12 features. Each epoch was linearly detrended before Welch power-spectrum estimation with `nperseg=256` and density scaling. The frozen SciPy implementation used a periodic Hann window and constant segment detrending; each 256-sample epoch therefore supplied one full-length spectral segment [14]. For each channel, relative power was calculated in delta [1,4), theta [4,8), alpha [8,13), beta [13,30), and gamma [30,45) Hz, together with a theta/beta ratio. Trapezoidal integration used the discrete bins selected by each half-open band mask, divided by integrated power over the inclusive 1–45 Hz mask.

Denominators were bounded below by machine epsilon. The 0.5 Hz frequency grid and the selected trapezoidal-integration endpoints are documented in S2 File. These discrete masks define the implemented band powers; the nominal frequency ranges should not be interpreted as a different, continuous integration procedure.

The amplitude-derived set contained six features: peak-to-peak range, mean absolute value, and population standard deviation for each channel, calculated before spectral detrending. The combined set concatenated all 18 features. The amplitude set was a simple feature comparator, not a disease-free negative control. These feature sets were not assumed to encode mutually exclusive physiological or acquisition effects. Participant identifiers, filenames, and sex labels were not classifier inputs.

Logistic regression (LR) used balanced class weights, C=1, the lbfgs solver, and a maximum of 3,000 iterations. Random forest (RF) used 300 trees, `balanced_subsample` class weights, square-root feature subsampling, and one worker per fit. Other parameters followed the frozen implementation defaults. StandardScaler transformations were fitted on training rows only, for both classifiers. Equal primary epoch counts gave participants equal numbers of training rows; this equality does not extend to every source-budget sensitivity analysis.

### Evaluation and participant-level metrics

Internal evaluation used five-fold StratifiedGroupKFold with shuffling and seeds 42–46, yielding five repetitions [15]. Within a repetition, that seed was used for both fold shuffling and the classifier random state. All epochs from a participant remained in one fold. The scaler and classifier were fitted separately in each training fold. The model’s `predict_proba` output for the ADHD class was averaged across each held-out participant’s epochs, yielding one out-of-fold (OOF) score per participant per repetition. A fixed threshold of 0.5 generated class predictions. The reported estimate is the mean of the five complete repetition-level metrics, not a metric computed after averaging scores across repetitions. The repetitions reuse the same participants and are not independent cohorts.

For each transfer direction, the scaler and classifier were fitted using all selected source epochs with model seed 42 and applied unchanged to the other cohort. Target labels were used for evaluation, not transformation fitting, hyperparameter tuning, threshold selection, or score reversal within that run. Internal and transfer evaluations therefore use the same pipeline specification but different fitted models and evaluation samples. Their juxtaposition is descriptive; no paired test of an internal-to-transfer performance difference was performed.

ADHD was encoded as the positive class. BA was the mean of sensitivity and specificity. AUC evaluated participant-level ADHD scores in the source-defined orientation, with tied case–control scores contributing one half to the ranking statistic. BA assesses decisions at the fixed 0.5 threshold, whereas AUC assesses ranking.

Probability outputs were treated as model scores; calibration and clinical decision utility were not evaluated. Accuracy and confusion matrices were retained in the aggregate record.

### Uncertainty and permutation analyses

We used 2,000 diagnosis-stratified participant bootstrap resamples, reporting the 2.5th and 97.5th percentiles as 95% conditional intervals. Transfer intervals condition on the fitted source model and stored target scores. For internal evaluation, the same sampled participant indices were applied to all five OOF repetitions before their metrics were averaged. Spectral-minus-amplitude differences used paired resamples and identical outer splits. These descriptive intervals exclude model refitting and do not establish full repeated-sampling coverage for dependent OOF predictions. Neither a direct LR-versus-RF test nor a between-cohort test of the spectral-minus-amplitude difference was performed.

Three permutation procedures used participants, not epochs, as the label-exchange unit. **Internal label association** used 999 permutations for spectral LR, each rerunning the complete five-repetition, five-fold evaluation. The repetitions shared one permuted label vector, used consistently for training, stratification, and scoring. **Source-label refit transfer** used 999 source-label permutations, refitting spectral LR before evaluation against unchanged target labels. **Fixed-model target association** used 1,999 target-label permutations with fixed spectral LR or RF scores. The procedures respectively address within-cohort label association, transfer relative to models trained on arbitrary source labels, and target-label association of an existing score. Their p values are not interchangeable [16]. Exchanges were unrestricted within the relevant cohort and did not condition on sex, source, or other potential confounders.

For a statistic T, *p*_MC_=(b+1)/(B+1), where B is the number of sampled permutations and b counts null values satisfying *T*_null_ ≥ *T*_observed_ − 1e-12. Positive-tail tests used BA or AUC; the exploratory nondirectional test used |◻AUC◻−◻0.5◻| for both the observed and null statistics. Zero exceedances were reported at the finite plus-one resolution rather than as p=0 [17]. Holm adjustment was applied separately within each null-type/statistic family: two cohorts internally, two source-refit directions, or four fixed-target direction–model combinations. This does not adjust for all historical analysis choices. RF did not receive the internal or source-refit permutation procedure, and spectral p values were not assigned to other feature sets. S2 File reports the 24 tests, exceedance counts, correction families, and Monte Carlo intervals for tail probabilities; those intervals are distinct from performance intervals.

### Sensitivity analyses and computational reproducibility

Sensitivity analyses replaced the first pediatric epochs with fifteen uniformly sampled epochs at seed 42; increased pediatric source caps to 31, 50, 80, or all available epochs while holding the adult target fixed; included all 80 adult records; and transferred models between the adult sex-defined subsets. A cap selected min(cap, available) epochs without excluding shorter records, changing both temporal coverage and potentially participant training weights. Historical 80-record/all-window settings were evaluated separately.

Analysis code, configuration, input hashes, and aggregate outputs were frozen together. The recorded environment was Python 3.13.5, NumPy 2.3.5, SciPy 1.17.0, and scikit-learn 1.8.0. The writing package contains aggregate results and table/figure code; participant-level data are still required for model refitting. Repeated generation of identical aggregate files documents computational reproducibility under recorded inputs, not biological reproducibility across new samples.

### Supporting evidence and AI-assisted workflow

A targeted comparison of 20 related records contextualizes the study; the search approach, access levels, and unresolved methods appear in S1 File. It is not an exhaustive systematic review. ChatGPT and OpenAI Codex (OpenAI) and Claude Code (Anthropic) assisted with code drafting, analysis checks, and figure integration; all reported values come from executed analyses and frozen outputs, and no plotted estimate was produced by an image-generation model. The author also used these tools to support literature discovery, organization, drafting, translation, and clarity checks of the manuscript text; the author reviewed and edited all content and takes full responsibility for it.

### Ethics statement

The study analyzed participant-level EEG recordings and supplied labels from existing public resources. The adult data provider reports informed consent and ethics committee approval for original data collection [13]. In email guidance, the University of California, Davis IRB Administration advised that this project would not constitute human subjects research and would not require IRB approval, provided that participants’ identities could not readily be ascertained from the information available to the researcher. This conditional guidance is not a formal determination issued through IRBNet. No new participant recruitment or clinical diagnostic adjudication was performed in this reanalysis. This was a retrospective analysis of archived data. The pediatric data were first accessed for research purposes on 8 July 2026 and the adult data on 9 July 2026. The analyzed files contain EEG signals, diagnostic group labels, recorded sex (adult cohort), and non-identifying record codes; the author had no access to information that could identify individual participants during or after data collection.

### Data and code availability

Adult EEG is available from Mendeley Data version 1, DOI 10.17632/6k4g25fhzg.1. The pediatric analysis used a supplied CSV mirror associated with IEEE DataPort DOI 10.21227/rzfh-zn36; the mirror identifier and unresolved upstream equivalence are documented in S2 File. Dataset access and reuse remain subject to the relevant distribution terms.

The accompanying computational materials contain frozen aggregate outputs, analysis configuration and hashes, model-analysis code, and table/figure generation code. Aggregate results are sufficient to rebuild the tables and figures; refitting requires the participant-level inputs described in the provenance record. The code and aggregate materials are archived as version v8 at https://doi.org/10.5281/zenodo.22858001; the archive carries the earlier working title of this study, and its frozen result files are byte-identical to those reported here (SHA-256 of the result matrix: 0c7c643d4ec4f18167116fc9cab6817b4c7a49ca82bc7098c8f3b67340ca6232). Original code is licensed under the MIT License; original documentation, figures, and aggregate outputs are licensed under CC BY 4.0. Third-party materials retain their original terms.

## Results

### Included records and data integrity

All 121 pediatric participants met the fifteen-epoch requirement. Adult channel screening retained 79 of 80 records. Three adult task-cell pairs—cell2/cell8, cell3/cell9, and cell4/cell10—had identical shapes, data types, and decoded sample bytes across all 80 records, giving 240 identical record-pair arrays. Because the primary experiment used cell1 only, these later duplicates did not increase its participant or epoch counts. The available files do not establish where in acquisition or data handling the duplication arose.

### Within-cohort discrimination and feature comparisons

Adult spectral LR and RF reached mean BA=0.9519 and 0.9546, with AUC=0.9883 and 0.9855, respectively (Table 1; Fig 2). Their conditional BA intervals were [0.9016, 0.9892] and [0.9059, 0.9919]. Pediatric spectral estimates were lower under the same two-channel specification: BA=0.5504 and 0.5664, with AUC=0.5665 and 0.5836. Combined pediatric features produced BA=0.5911 for LR and 0.6161 for RF; the RF AUC was 0.6842. Full internal results, including AUC intervals, appear in Table S2 in S2 File and Fig S1 in S2 File.

**Fig 2.**
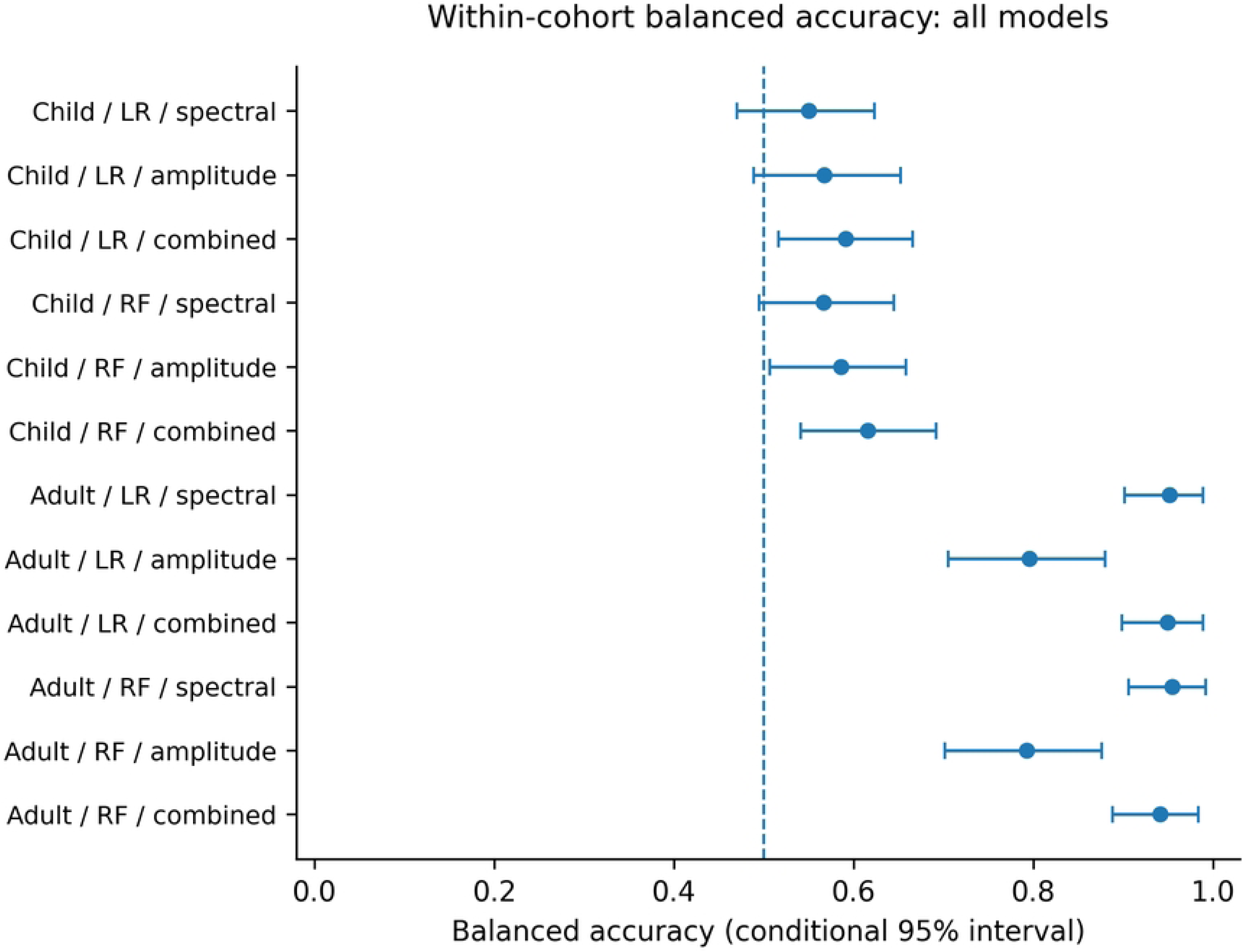
Within-cohort balanced accuracy for each classifier and feature set. Points are means of five repetition-level participant metrics. Horizontal bars are 95% conditional percentile intervals from diagnosis-stratified participant resampling of stored out-of-fold predictions; they omit full model-refitting uncertainty. The dashed line marks 0.5. Child: n=121; adult: n=79. AUC and paired feature differences are shown in Figs S1 and S2 in S2 File, respectively. LR, logistic regression; RF, random forest. The repetitions reuse the same participants.

**Table 1.**
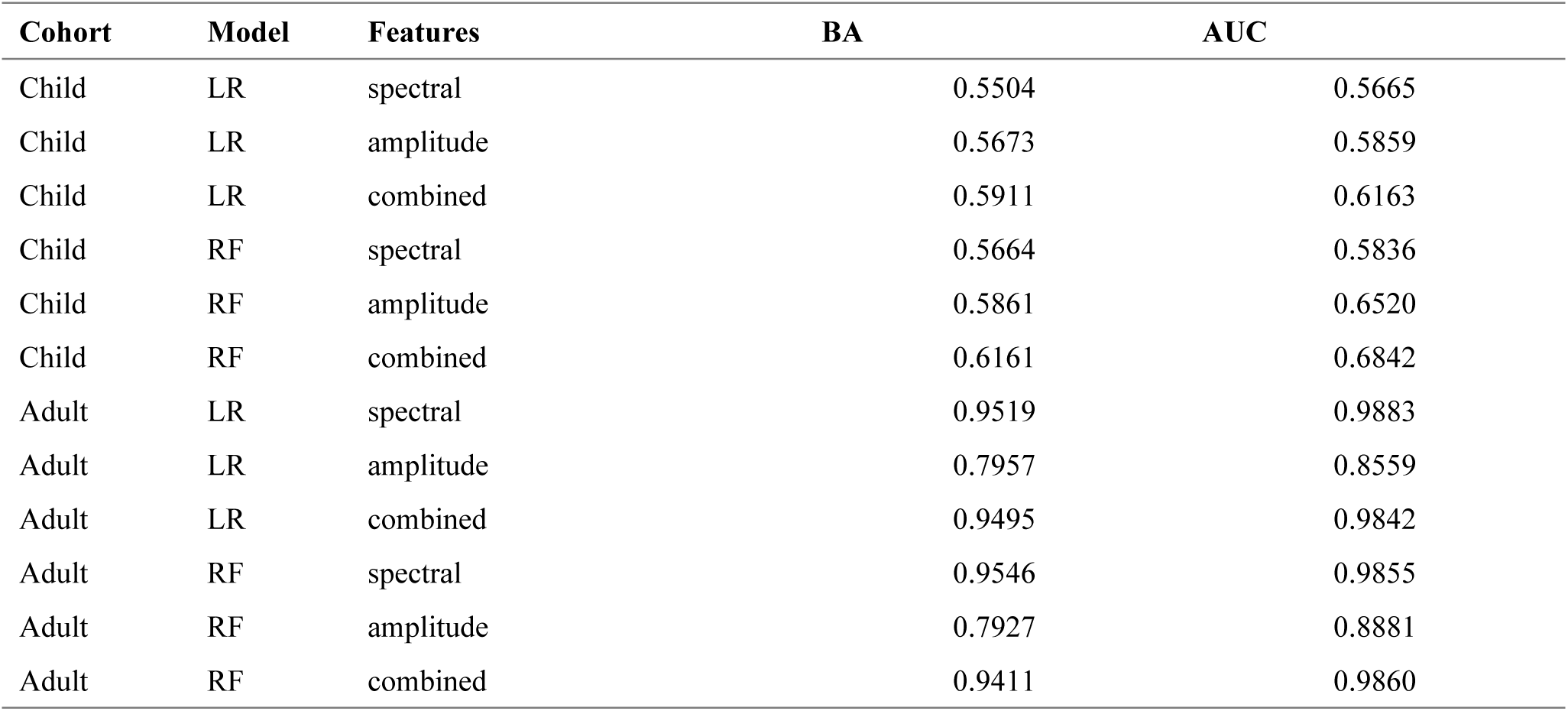
Within-cohort participant-level performance. Values are means of five repetition-level metrics. Child: n=121; adult: n=79. LR, logistic regression; RF, random forest; BA, balanced accuracy; AUC, area under the receiver operating characteristic curve. Conditional intervals appear in Table S2 in S2 File.

| Cohort | Model | Features | BA | AUC |
| --- | --- | --- | --- | --- |
| Child | LR | spectral | 0.5504 | 0.5665 |
| Child | LR | amplitude | 0.5673 | 0.5859 |
| Child | LR | combined | 0.5911 | 0.6163 |
| Child | RF | spectral | 0.5664 | 0.5836 |
| Child | RF | amplitude | 0.5861 | 0.6520 |
| Child | RF | combined | 0.6161 | 0.6842 |
| Adult | LR | spectral | 0.9519 | 0.9883 |
| Adult | LR | amplitude | 0.7957 | 0.8559 |
| Adult | LR | combined | 0.9495 | 0.9842 |
| Adult | RF | spectral | 0.9546 | 0.9855 |
| Adult | RF | amplitude | 0.7927 | 0.8881 |
| Adult | RF | combined | 0.9411 | 0.9860 |

The adult spectral-minus-amplitude BA differences were +0.1562 for LR and +0.1619 for RF; both paired conditional intervals were positive (Table 2). Pediatric amplitude BA estimates were slightly higher than the spectral estimates, but both paired intervals spanned zero. These comparisons condition on stored OOF predictions; they do not establish full training-procedure uncertainty or test whether the feature difference varies between cohorts. Fig S2 in S2 File displays the paired differences and intervals.

**Table 2.** Spectral-minus-amplitude differences on identical outer splits. Positive values favor the spectral model. The 95% percentile intervals use paired participant resamples of stored OOF predictions; they exclude model-refitting uncertainty. LR, logistic regression; RF, random forest.

| Cohort | Model | ΔBA | 95% conditional interval | ΔAUC | 95% conditional interval |
| --- | --- | --- | --- | --- | --- |
| Child | LR | -0.0169 | [-0.1172, 0.0814] | -0.0194 | [-0.1379, 0.1007] |
| Child | RF | -0.0198 | [-0.1122, 0.0732] | -0.0684 | [-0.1749, 0.0424] |
| Adult | LR | 0.1562 | [0.0710, 0.2425] | 0.1324 | [0.0534, 0.2277] |
| Adult | RF | 0.1619 | [0.0815, 0.2479] | 0.0974 | [0.0347, 0.1727] |

For adult spectral LR, no internal permutation reached the observed BA or AUC: both positive-tail $p_{MC}$ values were 0.001, with within-family Holm-adjusted values of 0.002. Pediatric spectral LR had 139 BA and 129 AUC exceedances among 999 permutations, corresponding to $p_{MC}$ =0.140 and 0.130. The adult result supports label association for the specified pipeline under the exchangeability null. The pediatric non-rejection is not evidence of equivalence to chance; neither result supplies an unperformed test for RF or other feature sets.

### Fixed-direction cross-cohort transfer

All four spectral transfer AUC estimates and their conditional intervals lay below 0.5 (Table 3; Fig 3). Pediatric-to-adult LR gave BA=0.3633 and AUC=0.0309; RF gave BA=0.3584 and AUC=0.2503. Adult-to-pediatric LR and RF gave BA=0.4593 and 0.4675, with AUC=0.3749 and 0.3857. The adult-to-pediatric BA intervals narrowly spanned 0.5, unlike their AUC intervals. Low AUC here describes reverse ranking in the fixed source-defined direction. At the unchanged threshold, pediatric-to-adult spectral LR had sensitivity 0.7027 and specificity 0.0238. The aggregate confusion counts for every transfer configuration are given in Table S4c in S2 File, alongside the distinct ranking and threshold metrics in S4a–b.

**Fig 3.**
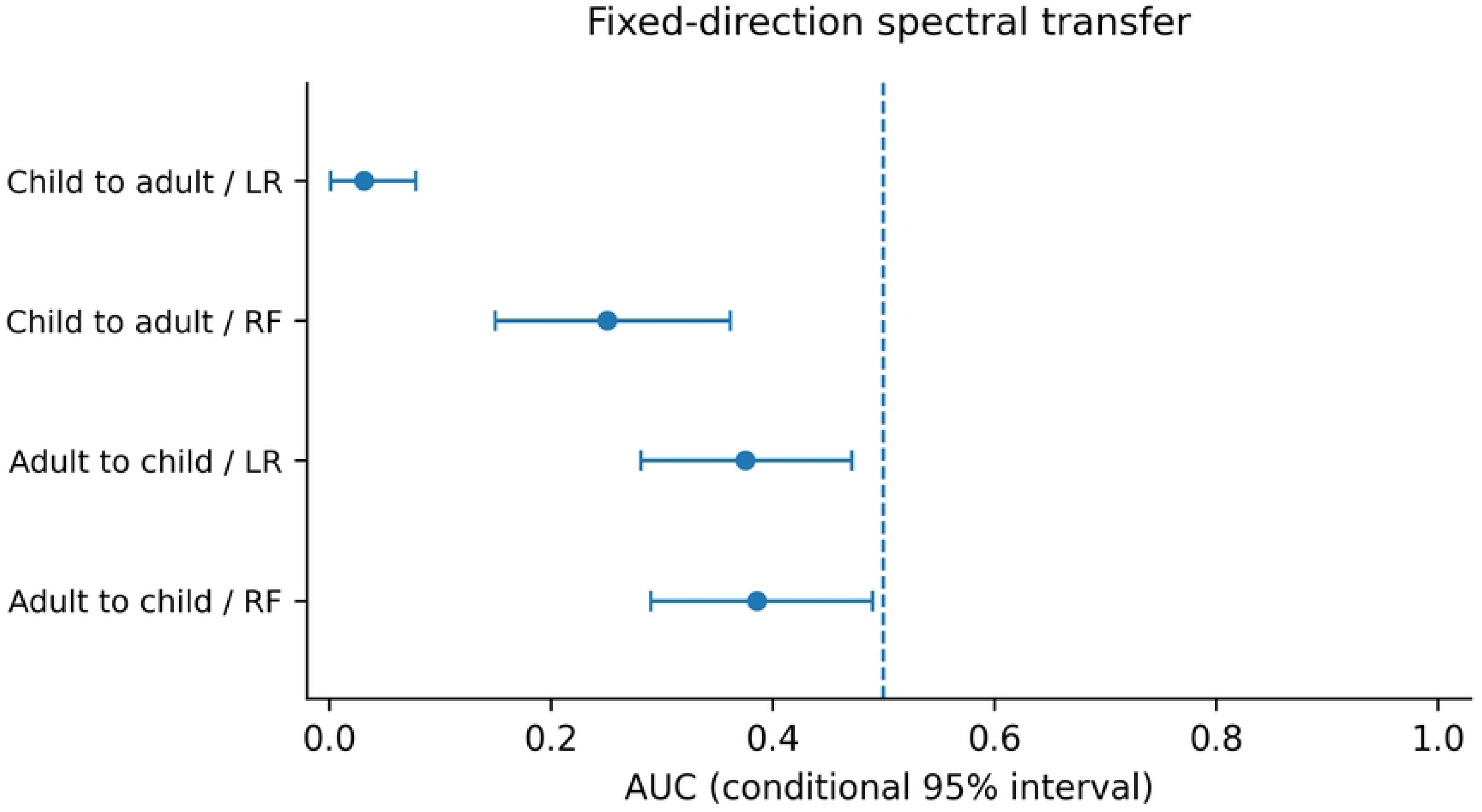
Spectral transfer AUC in both directions. Scores retain their source-defined ADHD orientation. All four conditional intervals lie below the 0.5 reference. Intervals condition on the fitted source model and stored target scores, not on new source-sample refitting. Corresponding BA estimates and intervals are reported separately in Table 3. LR, logistic regression; RF, random forest.

**Table 3.** Spectral transfer from source to target cohort, with score direction and the 0. 5 threshold unchanged. Child: n=121; adult: n=79. Intervals condition on the fitted source model and target scores. BA evaluates the threshold; AUC evaluates ranking. LR, logistic regression; RF, random forest.

| Direction | Model | BA | 95% conditional interval | AUC | 95% conditional interval |
| --- | --- | --- | --- | --- | --- |
| Child→Adult | LR | 0.3633 | [0.2838, 0.4411] | 0.0309 | [0.0013, 0.0779] |
| Child→Adult | RF | 0.3584 | [0.2654, 0.4498] | 0.2503 | [0.1499, 0.3617] |
| Adult→Child | LR | 0.4593 | [0.4178, 0.5007] | 0.3749 | [0.2809, 0.4716] |
| Adult→Child | RF | 0.4675 | [0.4262, 0.5089] | 0.3857 | [0.2904, 0.4903] |

For pediatric-to-adult spectral LR, the nondirectional AUC *p*_MC_ was 0.0005 under fixed-target label permutation (Holm-adjusted 0.002) and 0.020 under source-label refitting (19/999 exceedances; adjusted 0.040). The corresponding source-refit AUC null had standard deviation 0.2549 and central 95% quantiles [0.0521, 0.9466]. For adult-to-pediatric LR, fixed-target and source-refit nondirectional *p*_MC_ values were 0.0155 and 0.092. Its source-refit null standard deviation was approximately 0.077. Positive-tail spectral AUC tests did not provide evidence of positive transfer relative to their respective nulls. All 24 test results are reported in Table S7 in S2 File; matched-axis null-distribution plots are supplied as Fig S3a–d in S2 File.

### Source-budget and cohort-sensitivity results

With the 79-person adult target held fixed, pediatric-to-adult RF AUC increased from 0.2503 with fifteen source epochs per participant to 0.4897 with all available source epochs. LR AUC remained below 0.1 across the same budgets (Table 4; Fig 4). Uniform pediatric epoch sampling instead of first-epoch selection yielded transfer AUC=0.0907 for LR and 0.1293 for RF. In the reported budget series, LR remained strongly reverse-ranked while RF moved toward, but not above, an AUC of 0.5.

**Fig 4.**
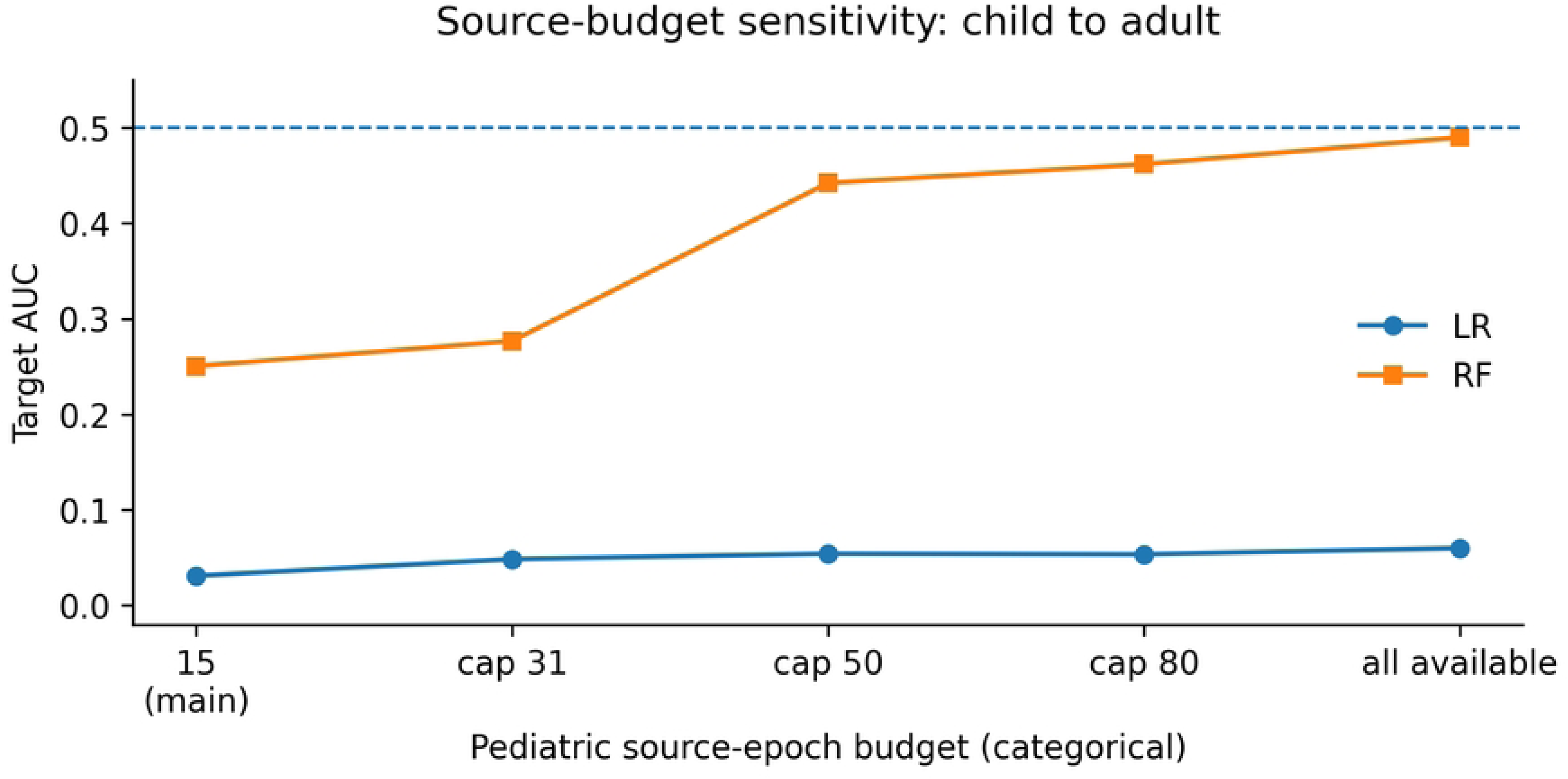
Pediatric-to-adult spectral transfer AUC across source-epoch budgets. The adult target is fixed at 79 participants, cell1, first fifteen epochs. Each source cap selects min(cap, available) epochs per participant, changing temporal coverage and potentially training weights. LR remains below 0.1; RF approaches 0.5. Lines connect the reported discrete specifications and do not represent interpolation to untested budgets. LR, logistic regression; RF, random forest.

**Table 4.** Pediatric-to-adult spectral transfer as the source-epoch cap changes. The adult target remains the 79-participant cell1 cohort with fifteen epochs per person. A cap selects min(cap, available); All uses every available source epoch. The models are refitted at each cap, with the remaining specification unchanged. LR, logistic regression; RF, random forest.

| Source cap | LR BA | LR AUC | RF BA | RF AUC |
| --- | --- | --- | --- | --- |
| 15 | 0.3633 | 0.0309 | 0.3584 | 0.2503 |
| 31 | 0.3784 | 0.0483 | 0.3990 | 0.2767 |
| 50 | 0.3919 | 0.0541 | 0.4720 | 0.4421 |
| 80 | 0.3919 | 0.0534 | 0.4887 | 0.4617 |
| All | 0.4324 | 0.0598 | 0.5174 | 0.4897 |

### Transfer across adult sex-defined subsets

Training on 23 female participants and evaluating on 56 male participants yielded spectral LR BA=0.9655/AUC=0.9911 and RF BA=0.9642/AUC=0.9923. In the reverse direction, both models had BA=0.8615, with AUC=0.9846 for LR and 0.9769 for RF. The association was observed in both transfer directions between sex-defined subsets of the same dataset. The directions also used different source and target sample sizes, so their performance difference does not isolate a sex effect; both retain the same acquisition source. Tables S6a and S6b in S2 File report every feature combination, source/target class count, and corresponding conditional BA and AUC interval.

## Discussion

### Internal discrimination and transfer describe different properties

The adult cohort supported strong ADHD-label discrimination after basic channel screening and participant-grouped validation. The internal LR permutation analysis supported this association, and performance remained high across the adult sex-defined subsets. In contrast, source-fitted spectral scores ranked cases below controls in both cross-cohort directions. These observations distinguish prediction within the sampled cohort from performance of a source-fitted rule in another population and task. They do not estimate a paired decline of one identical fitted model or identify what caused the cross-cohort difference.

The pediatric findings prevent a symmetric interpretation of this contrast. Spectral models were comparatively weak under the two-channel, thirty-second specification, while combined features yielded higher descriptive performance. The study therefore does not show two equally strong internal classifiers both losing their accuracy after transfer. Likewise, adult spectral-minus-amplitude differences estimate a predictive comparison, not a separable ADHD-specific biological component. Positive adult intervals and pediatric intervals spanning zero do not themselves test a between-cohort interaction. Neither the amplitude baseline nor the feature comparison identifies the cause of diagnostic-label association.

### Interpreting reverse ranking and specification sensitivity

An AUC below 0.5 indicates that target cases tend to receive lower scores than controls in the source-defined direction. It does not imply that scores contain no label information or that reversing them would generalize to another sample. Reversal chosen after inspecting target labels would be a target-informed analysis; the present runs retained their source-defined orientation. Some comparator models assigned every target participant to the same class at the fixed threshold despite different score rankings. Reporting BA and AUC separately exposes that distinction. Such output does not establish whether recalibration or a different threshold would improve performance: neither intervention was evaluated here. A degenerate BA interval describes resampling of fixed class assignments, not certainty about future classification performance.

Common channels, sampling rate, and duration did not align the measurement contexts. With one pediatric visual-task cohort and one adult resting-state cohort, population, task, and dataset are inseparable in this comparison. Developmental stage, recruitment, reference conventions, and voltage calibration remain additional plausible contributors [8, 9]. The design therefore characterizes the joint cross-cohort change rather than identifying an age effect, a task effect, or a unique acquisition artifact. Increasing the source-epoch cap also changes temporal coverage and, for records of different lengths, training weights. Its effect cannot be attributed to sample count alone. RF approached an AUC of 0.5 with more source epochs, whereas LR remained below 0.1 across the listed budgets. The magnitude of transfer therefore depended on classifier and data use, without evidence from this budget series that every model repeatedly changed direction.

The diagnosis-defined file grouping is a limitation of the available provenance, not evidence by itself that recording or preprocessing differed between diagnostic groups. The pediatric-to-adult source-label refit distribution was broad (AUC standard deviation 0.2549; central 95% quantiles 0.0521–0.9466), indicating that models trained on randomized source labels could produce markedly different rankings in the same adult target. A strongly separable adult feature structure is one possible explanation, but this analysis did not establish its dimensionality or biological versus acquisition-related origin. The observed source-trained result nevertheless lay in the nondirectional tail of this distribution (Monte Carlo p=0.020; within-family Holm-adjusted p=0.040). Neither the broad null nor the tail result identifies a transfer mechanism. Cross-sex evaluation changes the sex-defined files being used, but remains within the same dataset and cannot exclude unmeasured differences that might align with diagnosis across those files.

### Inference, limitations, and data provenance

The permutation procedures condition on different objects. Internal permutation evaluates the full within-cohort statistic; fixed-target permutation evaluates an existing scoring rule; source-label refitting compares transfer with models trained on arbitrary source labels. A broad source-refit distribution is not a distribution of fits to newly recruited, correctly labelled source participants. Fixed-input repeatability, specification sensitivity, and generalization to new samples must therefore remain distinct.

The main limitations are the small, heterogeneous cohorts, limited clinical metadata, and prior inspection of target results. This secondary analysis did not evaluate prospective use in a clinical referral population; calibration and decision benefit were not assessed. The analysis is exploratory, and local Holm adjustments do not account for all historical choices. Conditional bootstrap intervals omit full model-refitting uncertainty and do not fully represent dependencies induced by overlapping CV training sets; small-sample validation uncertainty is a substantive concern in neuroimaging prediction [18]. Unrestricted permutations do not remove source or demographic confounding. Two channels and two simple classifiers do not represent the full pediatric montage or modern architectures. Basic signal-validity screening does not eliminate all ocular, muscular, or other artifacts. Diagnosis-defined file grouping leaves potential diagnosis-aligned handling differences unresolved, while adult cross-sex tests retain a common data source and a small female target group.

The pediatric mirror-to-epoch conversion was verified, but correspondence to the canonical raw files remains unresolved. The repeated adult task arrays are an observed property of the supplied files; they neither identify an acquisition mechanism nor affect the cell1-only experiment. These provenance limits accompany the analysis record rather than being treated as explanations established by model performance.

### Contribution and next validation step

Independent EEG–ADHD validation and evaluation-design audits already exist [1, 2, 3, 4, 6]. The contribution is a reproducible comparison that preserves several otherwise conflated outputs: within-cohort discrimination, target ranking, fixed-threshold classification, and sensitivity to source-epoch use. The adult spectral result, the constant-class outputs of some comparator models, and the classifier-specific budget responses illustrate why these outputs need separate reporting. The supplied full matrix and distinct permutation questions make the comparison inspectable without assigning a causal interpretation that the design cannot resolve.

A subsequent test with closer age and task alignment, an independently obtained target cohort, and prespecified adaptation rules would better distinguish competing explanations. Until then, the evidence concerns these fitted pipelines in these target cohorts, not a universal diagnostic limit or a unique confounding mechanism.

## Conclusions

In this exploratory two-cohort reanalysis, strong adult within-cohort ADHD-label discrimination coexisted with reverse cross-cohort ranking by the source-fitted spectral models. Feature comparisons and source-epoch sensitivity further qualified the pattern. A single internal score did not characterize performance in the other population and task. The results support reporting internal discrimination, target ranking, thresholded classification, and their uncertainty separately; they do not isolate the biological or acquisition-related cause of the cross-cohort difference.

## Data Availability

The data underlying this study are third-party public datasets that other researchers can access in the same manner as the author, without special privileges. The adult EEG data are available from Mendeley Data (version 1, https://doi.org/10.17632/6k4g25fhzg.1). The pediatric EEG data are available from IEEE DataPort (https://doi.org/10.21227/rzfh-zn36) the analyzed CSV copy is the Kaggle dataset “EEG Dataset for ADHD”, version 1 (https://www.kaggle.com/datasets/danizo/eeg-dataset-for-adhd/versions/1). All analysis code, the frozen configuration, input-file hashes, and the aggregate results needed to reproduce every table and figure are openly available from Zenodo (https://doi.org/10.5281/zenodo.22858001).

https://doi.org/10.21227/rzfh-zn36

https://doi.org/10.17632/6k4g25fhzg.1

https://doi.org/10.5281/zenodo.22858001

## Supporting information

**S1 File. Related-work evidence register.** Targeted comparison of 20 related records, search and access record, and design distinctions.

**S2 File. Supplementary methods, aggregate results, and supplementary figures.** Tables S1–S9 and Figs S1– S3.

## Notes

### Competing Interest Statement

The authors have declared no competing interest.

### Author Declarations

University of California, Davis IRB Administration (email guidance, September 2026): this secondary analysis of existing, publicly available, de-identified EEG datasets would not constitute human subjects research and would not require IRB approval, provided that participants’ identities could not readily be ascertained by the researcher. This conditional guidance is not a formal IRBNet determination. The adult data provider reports informed consent and ethics committee approval for the original data collection.

